# An Integrated Large-Scale Atlas of Protein Quantitative Trait Loci across Olink and SomaScan platforms

**DOI:** 10.1101/2025.10.06.25336803

**Authors:** Chachrit Khunsriraksakul, Fan Zhang, Lida Wang, Siyuan Chen, Havell Markus, Dieyi Chen, Ganesh Shenoy, Carolina Lopez-Silva, Fayez Jabboure, Ayse E. Kesaf, Xiaowei Zhan, Joanna Melia, Ken Hui, Bibo Jiang, Dajiang J. Liu

## Abstract

Protein quantitative trait loci (pQTLs) provide insight into the genetic regulation of protein expression and disease biology. We performed a large-scale cross-platform meta-analysis of plasma proteomics, integrating data from over 90,000 individuals across Olink and SomaScan platforms. This effort identified >30,000 sentinel pQTLs, with multi-trait approach (MTAG) markedly boosting both discovery and replication, despite the potential differences in the protein measurements of the two platforms. While both platforms captured well-known pleiotropic loci (*HLA, ABO, SH2B3*), we also uncovered platform-specific associations, reflecting unique assay designs. We further established a comprehensive resource for transcriptome- and proteome-wide association studies (TWAS, PWAS), identifying >100,000 upstream regulators with strong replication. As a proof of concept, we applied this framework to inflammatory bowel disease (IBD), generating the largest to date GWAS directly comparing Crohn’s disease (CD) and ulcerative colitis (UC). Integrative multi-omics analyses revealed divergent loci enriched in the NF-*κ*B signaling pathway and improved CD vs. UC classification when proteomic features were combined with polygenic risk scores (PRS). Our findings provide a comprehensive resource for plasma protein genetics and demonstrate the value of integrative multi-omics for disease subtype classification, with broad implications for precision medicine.

## Introduction

Identifying protein quantitative trait loci (pQTL) is important for understanding the genetic basis of protein expression and its impact on various phenotypes. The advent of affinity-based proteomics has enabled classical assays to be performed in a multiplexed format with higher throughput and improved sensitivity compared to previous protein identification methods using mass spectrometry^1^. These advancements have made Olink, an antibody-based immunoassay, and SomaScan, an aptamer-based approach, prominent tools in pQTL analyses in large population cohorts.

To date, there have been several large-scale studies profiling both genetic information and plasma protein levels using Olink and SomaScan platforms^2-8^. Yet, little efforts exist to meta-analyze these datasets to maximize power. We present a comprehensive meta-analysis of pQTL data, encompassing over 46,000 European individuals with 2,941 Olink targets from the UK Biobank cohort and up to 45,000 European individuals with 5,880 SomaScan targets from the deCODE, AGES, INTERVAL, and KORA cohorts, with total sample sizes exceeding 90,000.

Previous comparisons between Olink and SomaScan platforms have been limited to relatively small cohorts^9-14^. In contrast, our meta-analysis leverages large-scale datasets from both platforms, enabling more robust cross-platform evaluation. We further integrated results across platforms to enhance discovery and improve diagnostic accuracy, and we established a comprehensive resource for transcriptome-wide association studies (TWAS) and proteome-wide association studies (PWAS) to support future research.

Proteomic data have already been shown to improve risk prediction across a range of common and rare diseases^15-21^. However, these efforts have largely focused on distinguishing patients with disease from healthy controls, rather than addressing diagnostic uncertainty within disease subtypes. As a proof of concept, we turned to inflammatory bowel disease (IBD), which encompasses Crohn’s disease (CD) and ulcerative colitis (UC). About 15% of IBD cases are difficult to classify because CD and UC can share overlapping symptoms, endoscopic features, and histopathology^22^. These uncertainties complicate clinical management, such as the role of colectomy, highlighting the need for better molecular tools to distinguish IBD subtypes. To address this, we used our newly developed proteomic resource in integrative multi-omics analyses to define molecular differences between CD and UC and built predictive models to distinguish the disease subtypes.

## RESULTS

### Overview of pQTL datasets

We utilized pQTL datasets profiled from large-scale proteomics platforms, including Olink and SomaScan. For Olink, we retrieved data from UK Biobank^13^, which consisted of 2,941 targets (mapped to 2,927 UniProt IDs) with a sample size of up to 46,218 individuals (**Figure 1A**). For SomaScan, we used data from four different cohorts (deCODE^6^, AGES^8^, INTERVAL^5^, KORA^4^), which probed 5,880 targets (mapped to 4,762 UniProt IDs) with a sample size of up to 45,225 individuals (**Figure 1A**). About 52.2% of SomaScan targets (3,074/5,880) were probed in at least three cohorts (**Figure 1B**).

**Figure 1.**
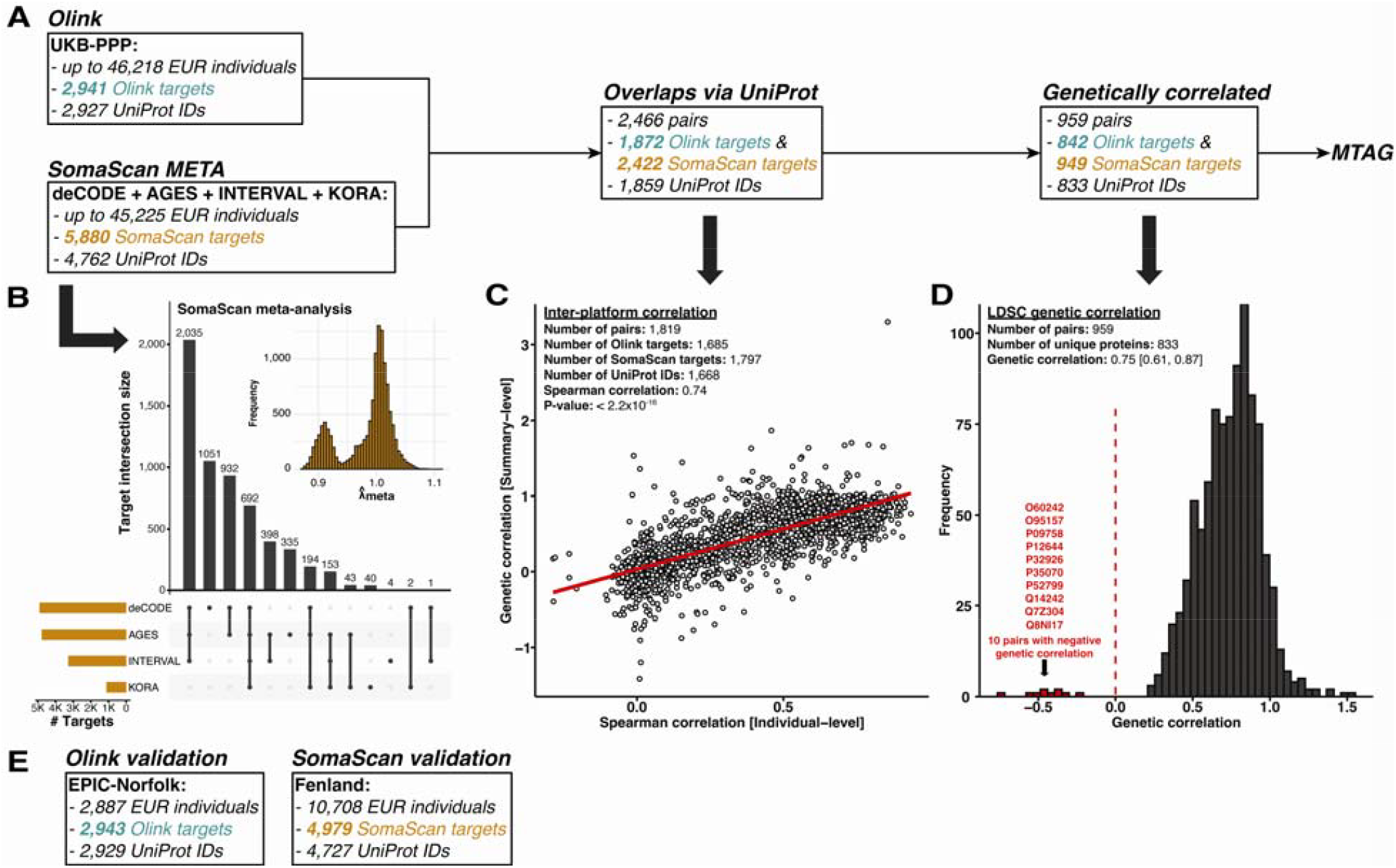
Cross-platform integration of Olink and SomaScan proteomic datasets. **(A)** Olink (UKB-PPP) and SomaScan (meta-analysis of deCODE, AGES, INTERVAL, and KORA) cohorts were analyzed. We consider overlapping targets mapped to the same UniProt ID as input. Proteins with significant genetic correlation were incorporated into MTAG analyses. **(B)** SomaScan data across four cohorts showed extensive overlapping targets and the meta-analyses have well-calibrated *λ*_*meta*_ statistics. **(C)** Inter-platform comparisons between the genetic correlation of pQTLs and phenotypic correlations of protein levels demonstrated strong concordance (Spearman correlation = 0.74). **(D)** LDSC genetic correlations of pQTL effects across 959 protein pairs (833 unique proteins) revealed a median genetic correlation of 0.75, with 10 pairs showing negative correlation. **(E)** Validation was performed in EPIC-Norfolk (Olink) and Fenland (SomaScan).

Prior studies reported modest correlation of protein measurements between the Olink and SomaScan platforms^13^. The protein correlation and genetic correlation may differ. To assess the genetic correlations between pQTLs, we assessed genetic correlations across overlapping proteins defined using UniProt IDs. This yielded 2,466 pairs, representing 1,872 Olink targets, 2,422 SomaScan targets, and 1,859 UniProt IDs (**Figure 1A**). For each pair of SomaScan and Olink proteins that share UniProtIDs, we calculate the genetic correlation between them. The median genetic correlation was 0.49 (interquartile range [0.12, 0.77]).

Next, we compared our genetic correlation estimates, derived from summary-level pQTL statistics, with Spearman correlations of protein levels, as calculated from individual-level Icelandic data^13^. Among 1,819 overlapping pairs, the correlation between the two approaches was 0.74 (P value < 2.2 × 10^-16^) (**Figure 1C**). Median genetic correlation was 0.51 [0.15, 0.78], while the median Spearman correlation was 0.42 [0.14, 0.63].

We then focused on protein pairs with significant genetic correlations between the Olink and SomaScan platforms, identifying 959 significant pairs representing 842 Olink targets, 949 SomaScan targets, and 833 UniProt IDs (**Figure 1A**). The median genetic correlation among these significant pairs was 0.75 [0.61, 0.87]. Notably, 10 pairs (e.g., BMP4, IL31RA) showed negative genetic correlations (**Figure 1D**). We subsequently applied multi-trait analysis using MTAG^23^ to meta-analyze pQTLs for targets with significant genetic correlations, leveraging genetic correlations across traits to enhance statistical power and thereby increase the discovery of pQTLs within each platform.

Lastly, we used the EPIC-Norfolk cohort (Olink; 2,943 targets; 2,887 individuals) and the Fenland cohort (SomaScan; 4,979 targets; 10,708 individuals) as independent replication cohorts to replicate identified pQTLs (**Figure 1E**).

### Discovery and Replication of pQTLs

We first evaluated the number of sentinel pQTLs across cohorts within each proteomic platform (**Figure 2A-B**). For Olink, the largest dataset came from the UKB-PPP cohort (N up to 46,218; 2,921 targets), which identified 17,378 sentinel pQTLs (2,173 *cis* and 15,205 *trans*) (**Supplementary Table 1**). In contrast, the smaller EPIC-Norfolk cohort (N = 2,887; 2,923 targets) identified 1,580 sentinel pQTLs (509 *cis* and 1,071 *trans*). For SomaScan, the deCODE cohort (N = 35,559; 4,907 targets) identified 16,221 sentinel pQTLs (1,888 *cis* and 14,333 *trans*), followed by FENLAND (N = 10,708; 4,979 targets) with 7,882 (1,602 *cis* and 6,280 *trans*), AGES (N = 5,368; 4,782 targets) with 3,448 (952 *cis* and 2,496 *trans*), INTERVAL (N = 3,301; 3,283 targets) with 1,831 (571 *cis* and 1,260 *trans*), and KORA (N = 997; 1,124 targets) with 232 sentinel pQTLs (169 *cis* and 63 *trans*).

**Figure 2.**
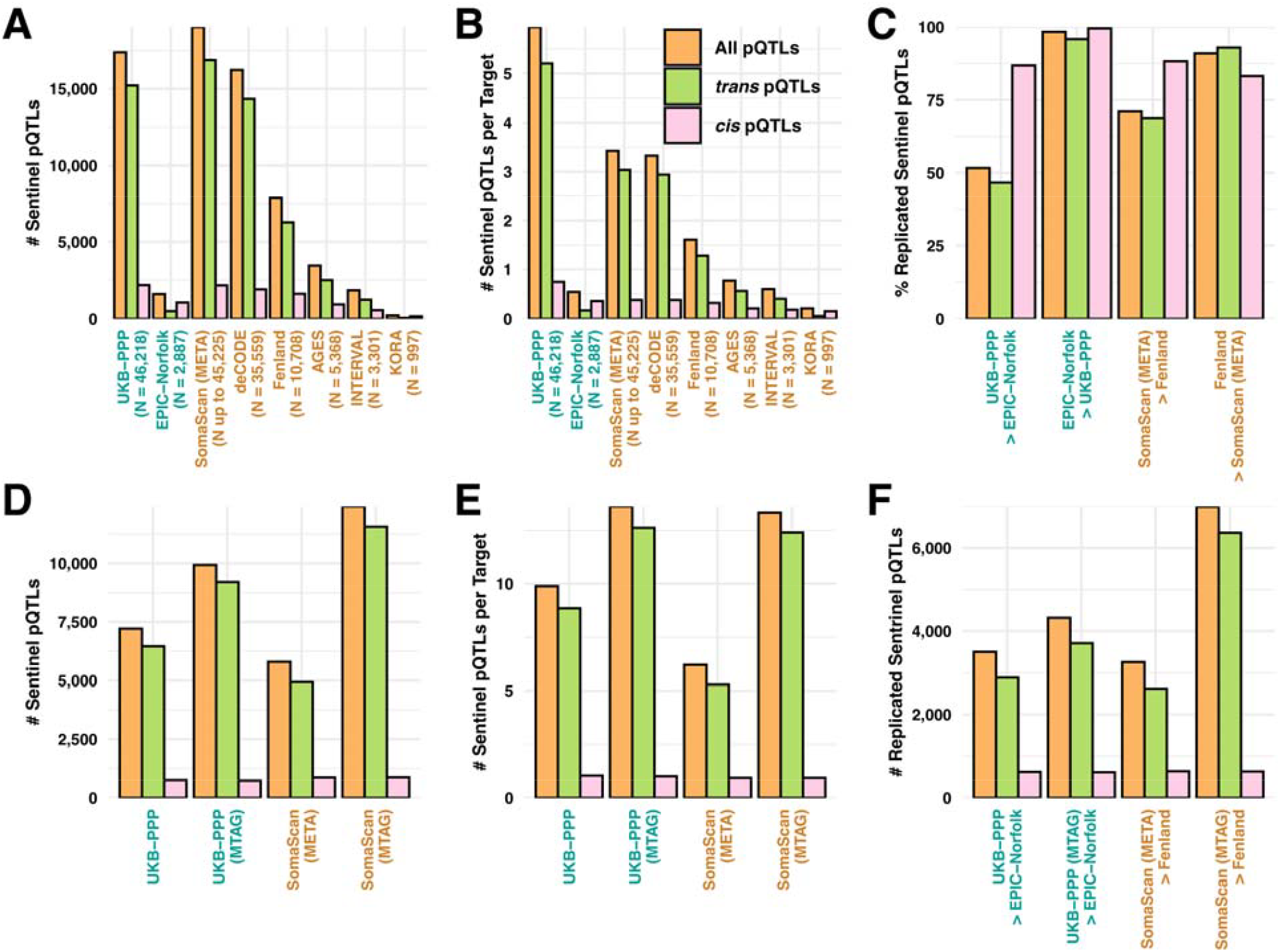
Discovery and replication of sentinel pQTLs across Olink and SomaScan platforms. **(A-B)** Number of sentinel pQTLs and sentinel pQTLs per target across cohorts, including UKB-PPP (Olink) and SomaScan (META, deCODE, Fenland, AGES, INTERVAL, and KORA). We stratify the numbers by *cis* and *trans* associations. **(C)** Replication rates of sentinel pQTLs in independent Olink (UKB-PPP and EPIC-Norfolk) and SomaScan (Fenland and META) datasets. **(D–E)** Number of sentinel pQTLs and sentinel pQTLs per target before and after multi-trait analysis (MTAG). **(F)** Replication of sentinel pQTLs after MTAG in Olink and SomaScan validation cohorts.

We next performed a pQTL fixed-effect meta-analysis across four cohorts (deCODE, AGES, INTERVAL, and KORA; N up to 45,225; 5,880 targets), referred to as SomaScan META. This analysis identified 19,030 sentinel pQTLs (2,158 *cis* and 16,872 *trans*) (**Supplementary Table 2**), representing a 17.3% increase in novel sentinel pQTLs compared to the largest SomaScan cohort analyzed to date. It is also worth noting that, on average, the Olink platform identified more sentinel pQTLs per target than SomaScan (5.9 vs. 3.4 pQTLs per target) (**Figure 2B**). To evaluate if the difference in the number of sentinel pQTLs per protein is due to sample size differences, we subsetted SomaScan datasets to sample sizes of at least 40K and 45K, making them comparable to Olink data in UK Biobank. In these comparable sample sizes, the numbers of sentinel pQTLs per target were 3.9 and 4.8, respectively for SomaScan. Given the similarity of the sample sizes of meta-analyzed Olink and SomaScan pQTL data, the difference in the average number of pQTLs per target likely reflects the measurement differences between the two platforms.

We subsequently evaluated the replication of pQTLs across cohorts (**Figure 2C**). We defined a pQTL as replicated if it has a P value < 0.05 and an effect direction consistent with the discovery sample. For the Olink platform, 86.8% of *cis* (1,692/1,949) and 46.7% of *trans* (6,451/13,813) associations identified in UKB-PPP were replicated in EPIC-Norfolk, whereas 99.6% of *cis* (893/897) and 95.9% of *trans* (417/435) associations identified in EPIC-Norfolk were replicated in UKB-PPP. For the SomaScan platform, 88.2% of *cis* (1,509/1,710) and 68.9% of *trans* (8,457/12,277) associations identified in SomaScan META were replicated in FENLAND, while 83.1% of *cis* (1,332/1,602) and 92.9% of *trans* (5,837/6,280) associations identified in FENLAND were replicated in SomaScan META. As expected, these findings demonstrate that replication rates are higher when smaller cohorts are validated in a larger cohort, underscoring the importance of the replication sample sizes for replication rates.

We then evaluated multi-trait meta-analysis of pQTLs, referred to as UKB-PPP (MTAG) and SomaScan (MTAG). Compared to single-cohort analyses, UKB-PPP (MTAG) identified 9,932 sentinel pQTLs, representing a 37.8% increase over UKB-PPP alone (7,210 sentinel pQTLs) (**Figure 2D**). The effect was even more pronounced for SomaScan, where SomaScan (MTAG) identified 12,439 sentinel pQTLs, a 114.6% increase compared to SomaScan (META) (5,796 sentinel pQTLs) (**Figure 2D**). Notably, the average number of sentinel pQTLs per target was highly comparable between UKB-PPP (MTAG) and SomaScan (MTAG), at 13.6 and 13.3 pQTLs per target, respectively (**Figure 2E**). MTAG analysis also substantially boosted replication, yielding a 23.3% increase in replicated sentinel pQTLs for the Olink platform (from 3,503 to 4,320) and more than doubling replication for the SomaScan platform, with a 114.7% increase (from 3,257 to 6,994) (**Figure 2F**).

We next examined pleiotropic loci captured by each platform (**Figure 3A, 3D**). Consistent with prior studies, we confirmed several well-established pleiotropic loci^2^, including *HLA, ABO, ZFPM2, ARHGEF3, SH2B3*, and *ASGR1*, all of which were detected by both Olink and SomaScan, underscoring strong cross-platform concordance. At the same time, we identified platform-specific pleiotropic signals, with Olink uniquely detecting loci such as *P4HA2, NRBF2, NFE2*, and *SERPINA1* (**Figure 3A**), while SomaScan uniquely captured *CFH, BCHE*, and *TMEM97* (**Figure 3D**). These findings highlight not only the overlap but also the distinct technical sensitivities of each platform.

**Figure 3.**
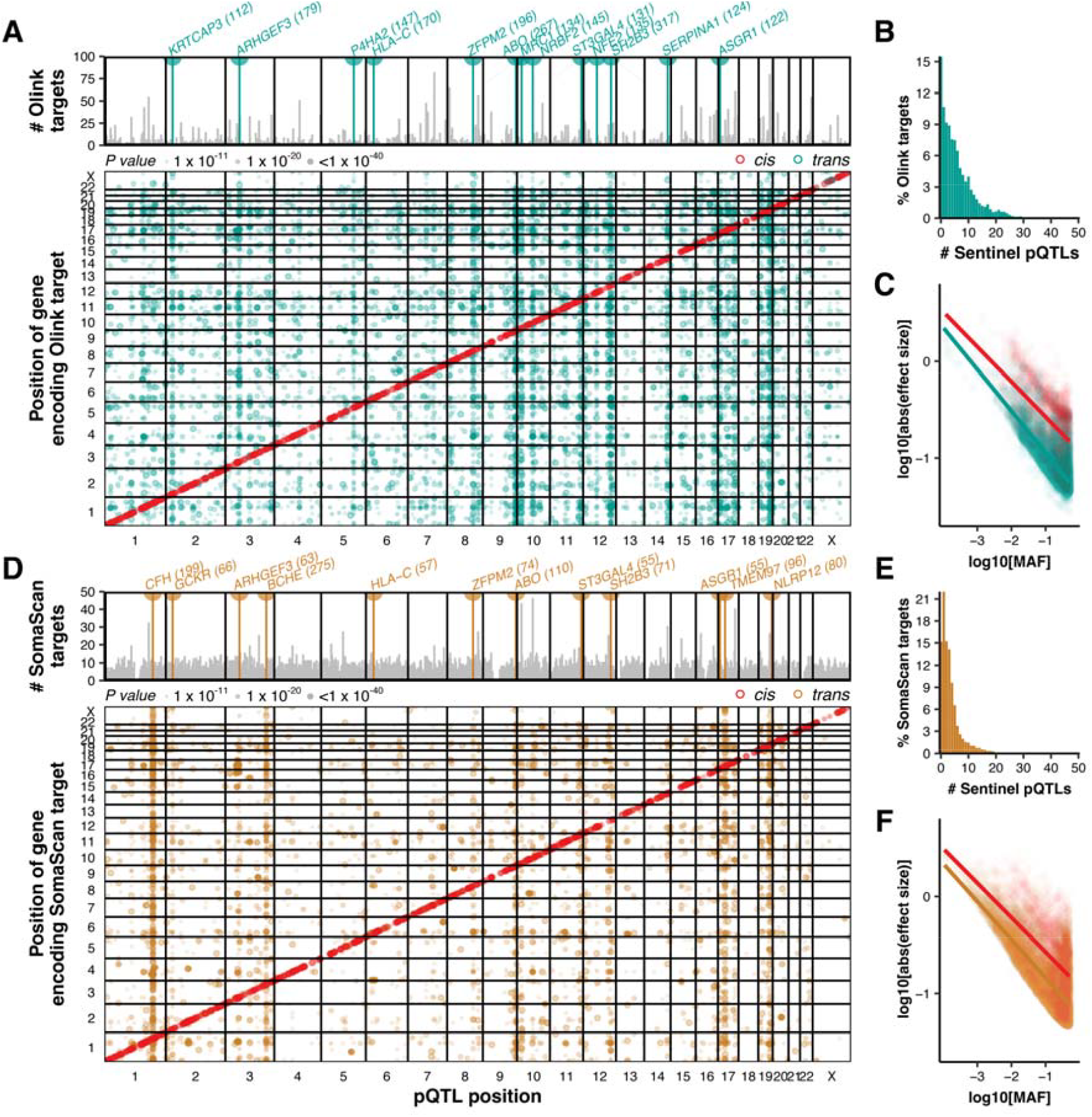
Genomic landscape of *cis*- and *trans*-pQTLs across Olink and SomaScan platforms. **(A)** Manhattan-style overview of sentinel pQTLs identified by Olink, with gene-encoding targets (Y-axis) and positions of sentinel variants (X-axis) highlighted, stratified by *cis* and *trans* associations. The top panel highlights pleiotropic loci. The y-axis shows the number of associated proteins, with the axis capped at 100. The regions with more than 100 associated proteins are labeled with counts in parentheses. **(B)** Distribution of Olink targets by number of sentinel pQTLs. (C) Relationship between minor allele frequency (MAF) and effect size for Olink sentinel pQTLs. **(D)** Sentinel pQTLs identified by SomaScan, with gene targets and variant positions shown, stratified into *cis* and *trans* associations. The top panel highlights pleiotropic loci. The y-axis shows the number of associated proteins. The axis is capped at 50 and regions with more than 50 associated proteins are labeled with counts in parentheses. **(E)** Distribution of SomaScan targets by number of sentinel pQTLs. **(F)** Relationship between MAF and effect size for SomaScan sentinel pQTLs.

For the Olink platform, we detected a median of 4 sentinel pQTLs per target (IQR: 1–9), with a maximum of 63 (**Figure 3B**). In contrast, the SomaScan platform showed a median of 2 sentinel pQTLs per target (IQR: 1–4), with a maximum of 33 (**Figure 3B**). Across both platforms, we observed a consistent inverse relationship between effect size magnitude and minor allele frequency (MAF) for both *cis* and *trans* associations, with *trans* associations generally exhibiting smaller effect sizes than *cis* associations (**Figure 3C, 3F**).

### Inferring tissue origins of proteins through pQTL enrichment analysis

Previous studies suggest that plasma proteins can be broadly divided into two main categories: highly abundant proteins predominantly synthesized by the liver, and a larger, more diverse set of lower-abundance proteins originating from various tissues and organs as part of normal cellular turnover^24,25^. To infer the tissue origins of plasma proteins, we conducted tissue enrichment analysis using LDSC-SEG using default settings. GTEx transcriptomic profiles across 53 tissues served as the reference dataset, enabling evaluation of enrichment patterns by assessing overlap between regions of tissue-specific expression and the pQTLs associated with each plasma protein. Across both the Olink and SomaScan platforms, our analyses identified the liver, small intestine, adipose tissue, spleen, and whole blood as the predominant contributors to the plasma proteome, whereas brain tissues contributed the least, likely due to the restrictive properties of the blood–brain barrier^26^ (**Figure 4A-B**).

**Figure 4.**
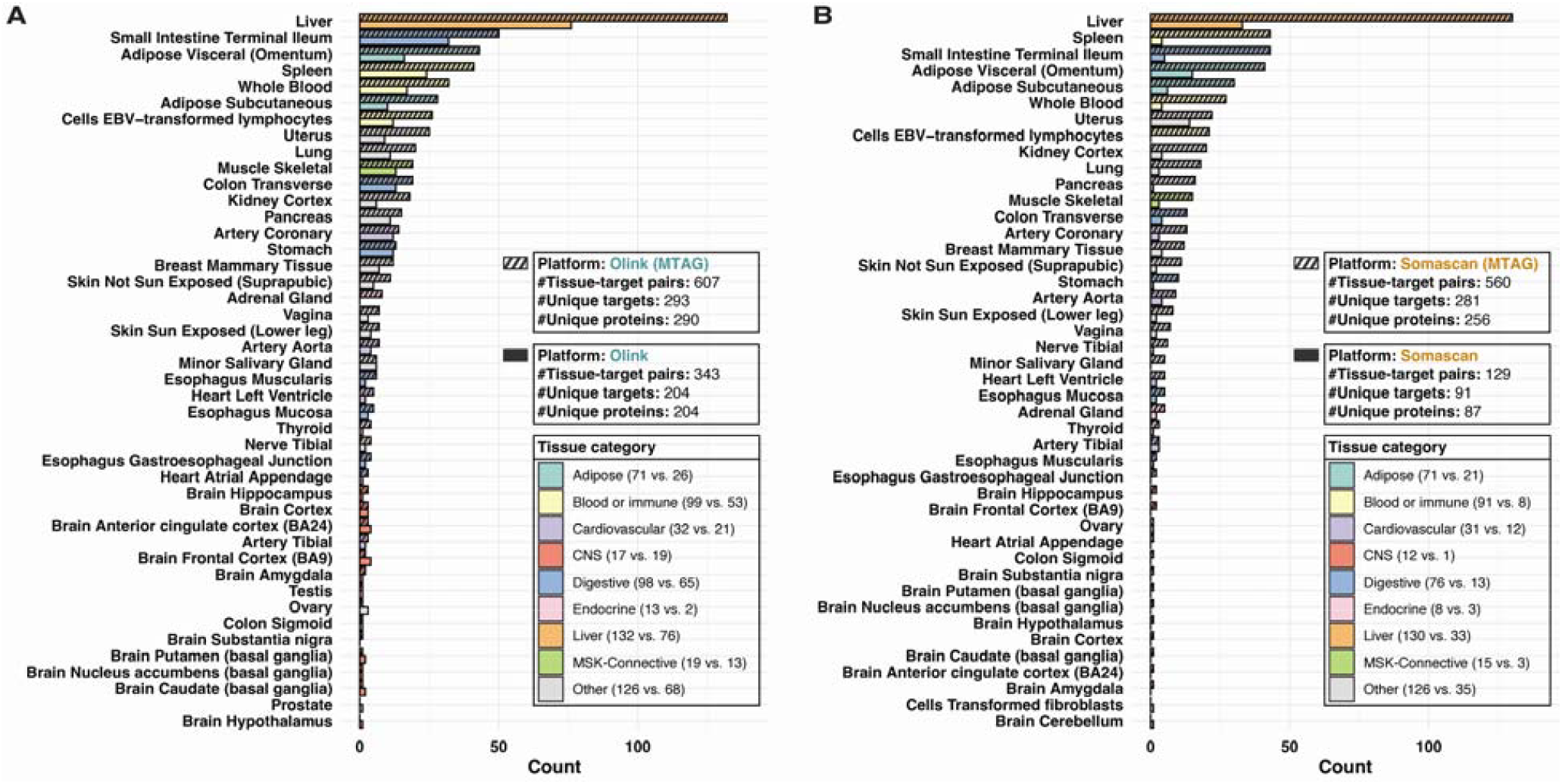
Tissue enrichment analysis of pQTLs across Olink and SomaScan platforms. **(A)** Olink platform. Bar plots depict the number of protein targets enriched in each GTEx tissue based on pQTL data. Enrichment analysis is done with LDSC with annotation tracks provided by LDSC-SEG. We compare results before MTAG (solid bars) and after MTAG (striped bars). Insets summarize total tissue-target pairs, unique targets, and unique proteins identified. **(B)** SomaScan platform. Results are similarly shown as in panel A. Tissue categories are color-coded (adipose, blood or immune, cardiovascular, central nervous system (CNS), digestive, endocrine, liver, musculoskeletal (MSK)-connective, and other). Across both platforms, protein targets are most enriched in liver, blood/immune, and adipose tissues, while brain tissues show the lowest overall enrichment.

### Transcriptome-wide association study of plasma protein levels

Using PUMICE software^27^, we performed TWAS of plasma protein levels to identify upstream regulators (**Figure 5A, 5D**). The top pleiotropic regulators included *GTF3C2, SLC22A5*, and *CYP21A1P* for Olink, and *CFHR1, SKIC2*, and *BCHE* for SomaScan (**Figure 5A, 5D**). Notably, *GTF3C2* is part of a core transcriptional machinery that broadly regulates transcription^28^. Upstream regulators were classified as *cis*-same, *cis*-different (depending on whether the *cis*-regulator is the same or different gene as the protein target), or *trans*, as previously described^29^ (**Methods**). For the UKB-PPP cohort, we identified 145,181 significant upstream regulators, including 1,669 *cis*-same, 28,557 *cis*-different, and 114,955 *trans* (**Figure 5B, Supplementary Table 3**). Notably, *cis*-same regulators were more broadly shared across tissues (median: 5 tissues) compared to *cis*-different (median: 2 tissues) or *trans* regulators (median: 2 tissues) (**Figure 5C**). Similarly, for the SomaScan META, we identified 149,302 significant upstream regulators, comprising 1,612 *cis*-same, 25,900 *cis*-different, and 121,790 *trans* (**Figure 5E, Supplementary Table 4**). As with Olink, *cis*-same regulators were more widely shared (median: 8 tissues) than *cis*-different (median: 2 tissues) or *trans* (median: 2 tissues) regulators (**Figure 5F**).

**Figure 5.**
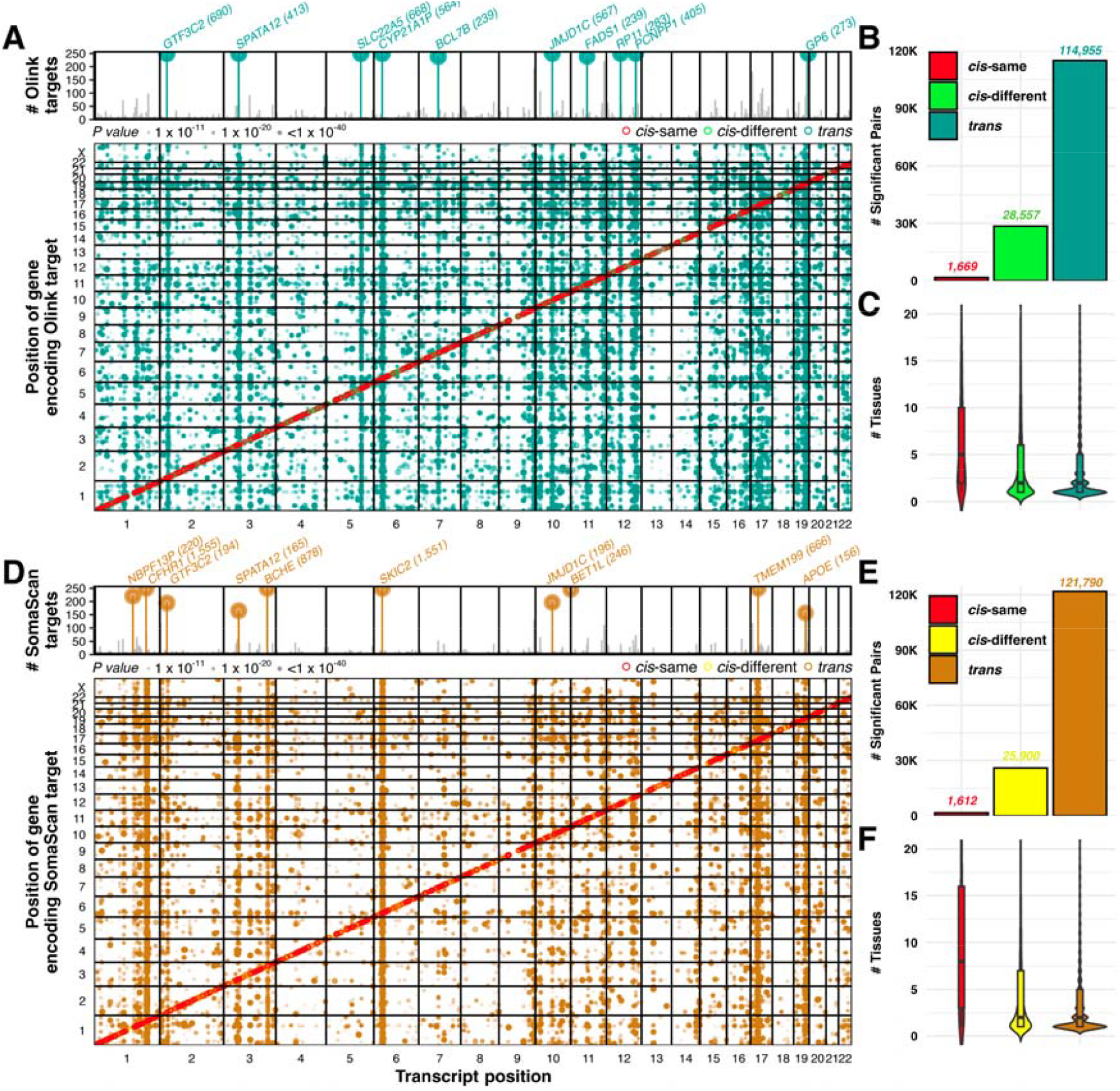
Genome-wide landscape of upstream regulators of plasma proteins across Olink and SomaScan platforms. **(A-C)** Olink results. **(A)** Genome-wide matrix of significant gene-protein associations from TWAS, with each dot representing the single most significant upstream regulator protein pair aggregated across all tissues. Dots are colored by association type: *cis-*same, *cis-*different, or *trans*, with the red diagonal marking *cis-*same relationships. The top panel highlights pleiotropic loci, upstream regulators influencing multiple protein targets, and emphasizes key regions with broad regulatory effects. The y axis represents the number of associated proteins and is capped at 250. The top 10 regulators are labeled with the number of associated proteins, as shown in parentheses. **(B)** Bar plot of the total number of significant gene-protein pairs, stratified by association type. **(C)** Violin plots showing the number of tissues in which each upstream regulator-protein pair is detected, stratified by association type. **(D-F)** SomaScan results. Panels show the same analyses as A-C for the SomaScan platform, summarizing the most significant upstream regulator for each protein across tissues and highlighting pleiotropic regulators with broad effects.

We next performed TWAS in the replication cohorts. We defined TWAS signals to be replicated if it has a P value < 0.05 and an effect direction consistent with the discovery sample. In EPIC-Norfolk, we identified 13,441 significant upstream regulators, including 893 *cis*-same, 8,257 *cis*-different, and 4,291 *trans* (**Figure 6A**). In Fenland, we identified 61,763 significant upstream regulators, comprising 1,343 *cis*-same, 16,493 *cis*-different, and 43,927 *trans* (**Figure 6A**). As expected, the replication cohorts yielded fewer regulators than the discovery analyses, reflecting their smaller sample sizes. Consistent with prior observations, the Olink platform identified more regulators per target than SomaScan (49.7 vs. 26.9 on average) (**Figure 6B**). We then assessed replication across cohorts (**Figure 6C**). For Olink, 80.0% of *cis*-same (1,336/1,669), 70.5% of *cis*-different (20,135/28,557), and 55.2% of *trans* (63,482/114,955) associations identified in UKB-PPP replicated in EPIC-Norfolk, whereas 82.9% of *cis*-same (740/893), 75.5% of *cis*-different (6,236/8,257), and 83.2% of *trans* (3,568/4,291) associations identified in EPIC-Norfolk replicated in UKB-PPP. For SomaScan, 83.9% of *cis*-same (1,352/1,612), 77.7% of *cis*-different (20,122/25,900), and 73.5% of *trans* (89,497/121,790) associations identified in SomaScan META replicated in Fenland, while 92.6% of *cis*-same (1,243/1,343), 88.7% of *cis*-different (14,635/16,493), and 73.5% of *trans* (37,935/43,927) associations identified in Fenland replicated in SomaScan META. Notably, replication rates were consistently higher for TWAS than pQTL analyses (Olink: 58.5% vs. 51.7%; SomaScan: 74.3% vs. 71.3%), showing that aggregating the effects of multiple genetic variants can improve the power for identifying regulatory effects for protein levels.

**Figure 6.**
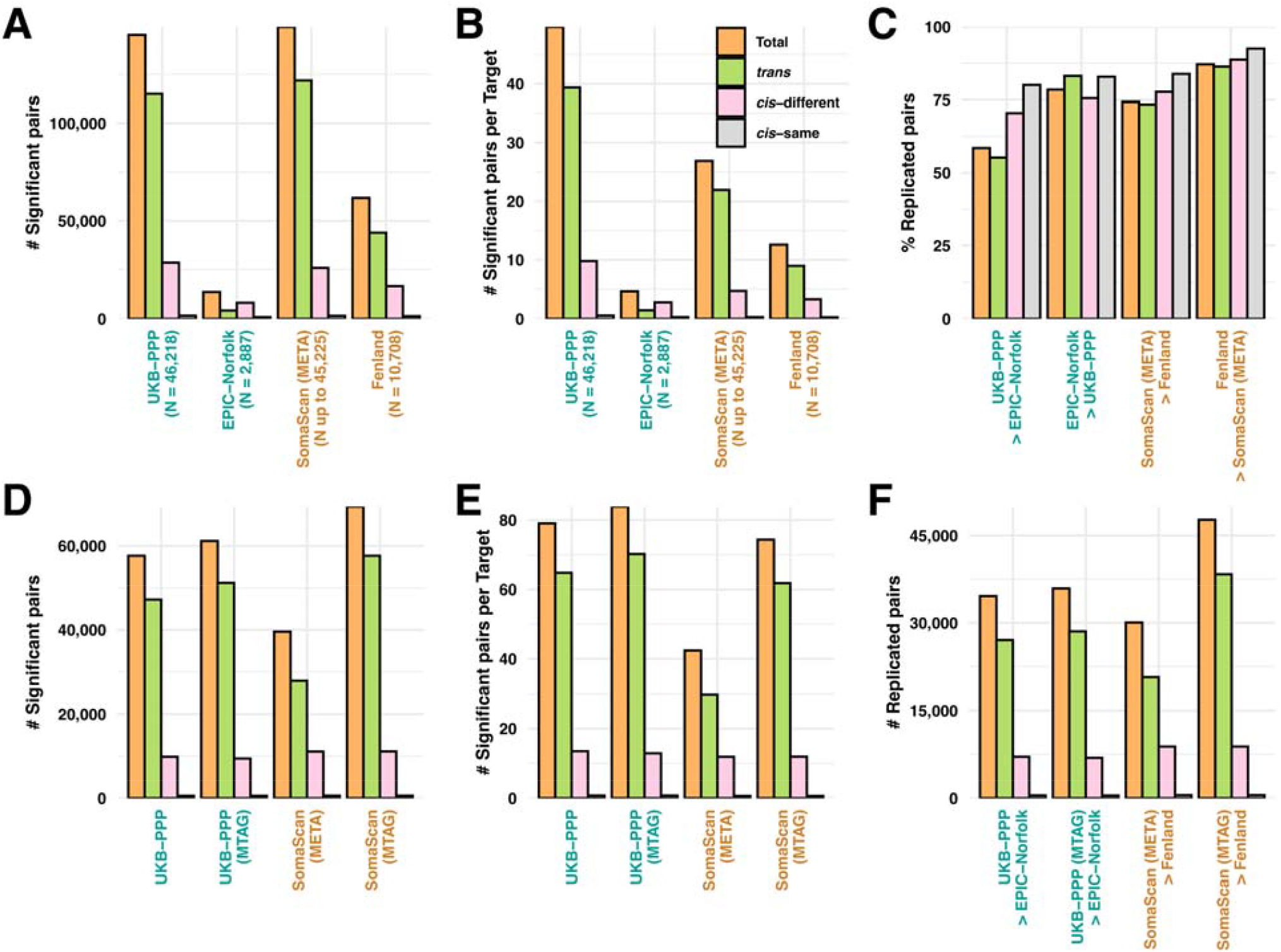
Discovery and replication of significant gene-protein pairs across Olink and SomaScan platforms. **(A)** Total number of significant gene-protein pairs identified in UKB-PPP, EPIC-Norfolk, SomaScan meta-analysis (META), and Fenland cohorts, stratified as *cis*-same, *cis*-different, and *trans*. **(B)** Number of significant gene-protein pairs per target across the cohorts. **(C)** Replication rates of significant pairs across independent Olink (UKB-PPP and EPIC-Norfolk) and SomaScan (META and Fenland) datasets. **(D)** Number of significant gene-protein pairs before and after MTAG in Olink (UKB-PPP) and SomaScan (META) datasets, categorized by association type. **(E)** Number of significant gene-protein pairs per target before and after MTAG. **(F)** Number of replicated gene-protein pairs following MTAG, highlighting increased discovery and replication across both proteomic platforms.

Lastly, we assessed the gain in TWAS signals when using MTAG datasets over the use of Olink or SomaScan data alone. In UKB-PPP (MTAG), we identified 61,151 significant regulators (572 *cis*-same, 9,402 *cis*-different, and 43,927 *trans*), compared with 57,625 regulators (1,343 *cis*-same, 16,493 *cis*-different, and 43,927 *trans*), representing a 6.1% increase (**Figure 6D**). For SomaScan (MTAG), we identified 69,378 regulators (625 *cis*-same, 11,103 *cis*-different, and 57,650 *trans*), compared with 39,552 regulators (625 *cis*-same, 11,093 *cis*-different, and 27,834 *trans*), corresponding to a 75.4% increase (**Figure 6D**). The number of regulators identified per target also increased with MTAG analyses (Olink: 83.9 vs. 79.0; SomaScan: 74.4 vs. 42.4) (**Figure 6E**). Importantly, MTAG boosted replication, resulting in a 3.7% increase in replicated sentinel pQTLs for Olink (from 34,641 to 35,940) and a 58.8% increase for SomaScan (from 30,071 to 47,739) (**Figure 6F**).

As a proof of concept, we investigated upstream regulatory program of NFKB1. Our TWAS analysis identified multiple candidate regulators with prior supporting evidence, including *SLC39A8* (*ZIP8*), *IRF1, CISD2, P4HA2, IL13, C4A, SLC22A4, SLC22A5, GPR84*, and *CLIC1*. In particular, *SLC39A8* (*ZIP8*) is a negative regulator of the NF-*κ*B signaling pathway through zinc-mediated suppression of IKK activity^30^. *IRF1* is required for full NF-*κ*B transcriptional activity at the long terminal repeat enhancer^31^. *CISD2* has been shown to antagonize the activation of NF-*κ*B by acting upstream of the peroxisome proliferator-activated receptor-*β*^32^. *P4HA2* activates the NF-*κ*B signaling pathway by promoting proteasome-dependent degradation of I*κ*B*α*^33^. IL13 was previously shown to induce an activation of NF-*κ*B which was demonstrated by the NF-*κ*B translocation to nuclei^34^. A larger *C4A* copy number is associated with enrichment of NF-*κ*B pathway^35^. *SLC22A4* and *SLC22A5* have been shown to play a role in the inflammatory process in Crohn’s disease, rheumatoid arthritis, and asthma^36-39^. *GPR84* agonist was previously found to upregulate NF-*κ*B signaling pathway and amplify the expression of inflammatory mediators^40^. Lastly, NF-*κ*B was positively regulated by *CLIC1*^41^.

### Development of protein imputation models

We built protein imputation models using PRS-CS^42^, a method previously applied to generate polygenic risk scores from summary statistics. For the Olink platform, we successfully imputed 1,712 of 2,941 protein targets, with a median prediction correlation of 0.41 (IQR 0.23–0.62). For the SomaScan platform, we imputed 2,528 of 4,972 targets, with a median correlation of 0.28 (IQR 0.17–0.47) (**Figure 7A**). We then evaluated the impact of incorporating MTAG datasets on the number of imputable models and prediction accuracy. The number of significant models increased modestly (Olink: 720 to 744; SomaScan: 693 to 732) (**Figure 7B**). In contrast, prediction accuracy improved significantly, with median correlation increasing by 6.3% (0.48 to 0.51, P value = 0.03) for Olink and by 20% (0.35 to 0.42, P value = 1.99 × 10^-9^) for SomaScan (**Figure 7C**).

**Figure 7.**
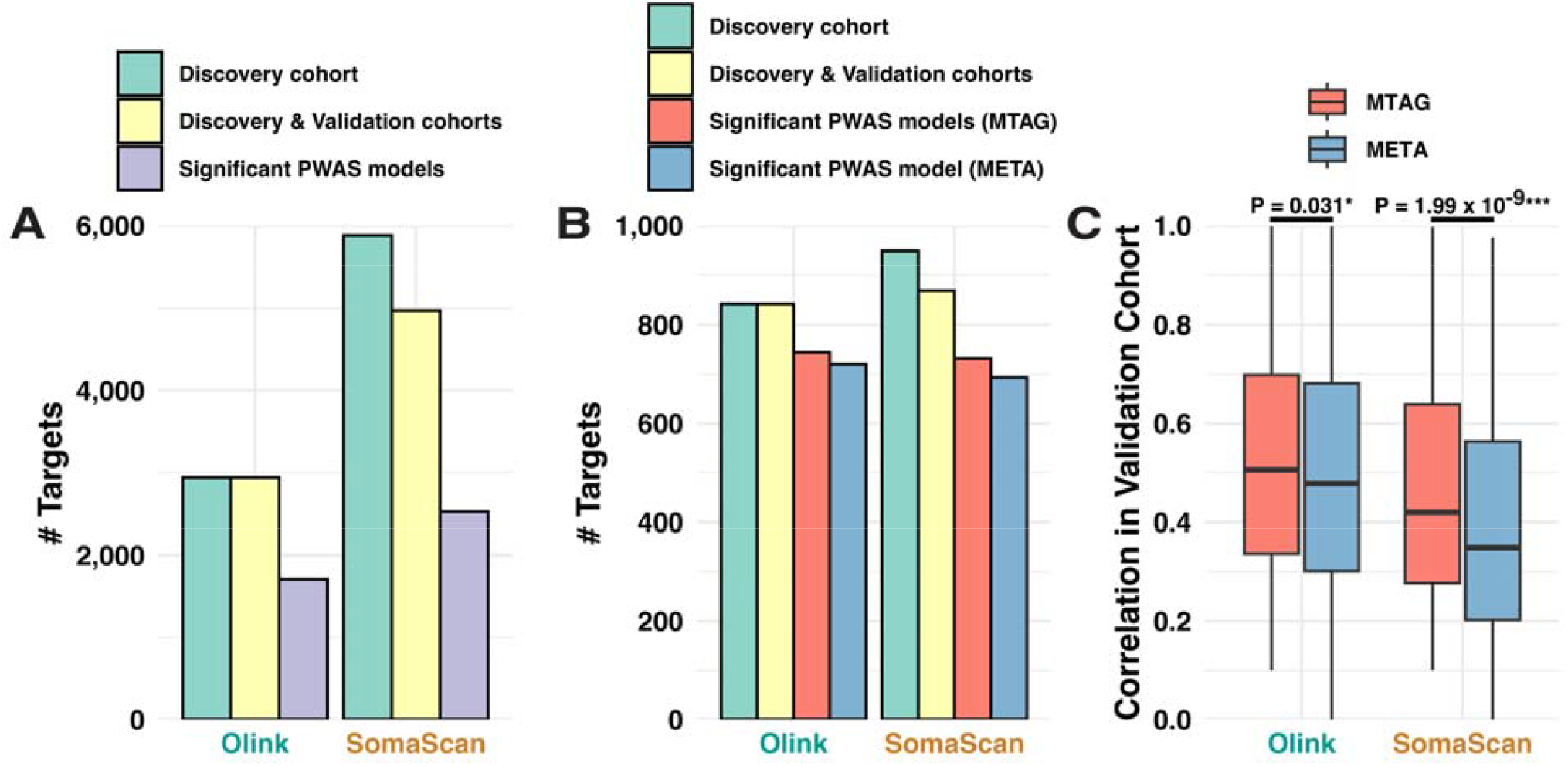
PWAS model development and validation across Olink and SomaScan platforms. **(A)** Number of protein targets profiled in the discovery cohort, jointly available in discovery and validation cohorts, and with significant PWAS models for Olink and SomaScan. **(B)** Number of targets with significant PWAS models derived using MTAG or standard meta-analysis (META) for each platform. **(C)** Boxplots showing prediction accuracy in independent validation cohorts for MTAG and META models. P values are derived from two-sided Mann-Whitney U test, comparing the accuracy difference between MTAG and META-based models.

### Imputation of transcriptome and proteome to identify divergent molecular regulators of CD and UC

We generated a novel GWAS dataset that directly compared 12,194 CD cases and 12,366 UC cases (**Figure 8A, Supplementary Table 5**) using CC-GWAS^43^. Subsequently, we performed downstream analyses, including TWAS and PWAS, to identify key differential molecular regulators (**Figure 8B-C, Supplementary Table 6-7**). Our findings revealed 15 divergent genetic loci through CD vs. UC GWAS, highlighting well-known genes such as *NOD2, HNF4A, PTGER3*, and *PTGER4* (**Figure 8A**). TWAS identified 27 novel loci, including *NFKB1* (**Figure 8B**), while PWAS uncovered 32 additional loci, such as CD8A, IL34, PLA2G10 and TFF3 (**Figure 8C**). Notably, several of the identified genes have previously been implicated in distinguishing CD from UC. For example, *HNF4A*, a regulator of mucosal barrier integrity, is downregulated in both CD and UC, with a more pronounced reduction in UC^44^. In addition, *NF-κB* levels are elevated in lamina propria biopsy specimens from patients with CD compared with those from UC patients and healthy controls^45^.

**Figure 8.**
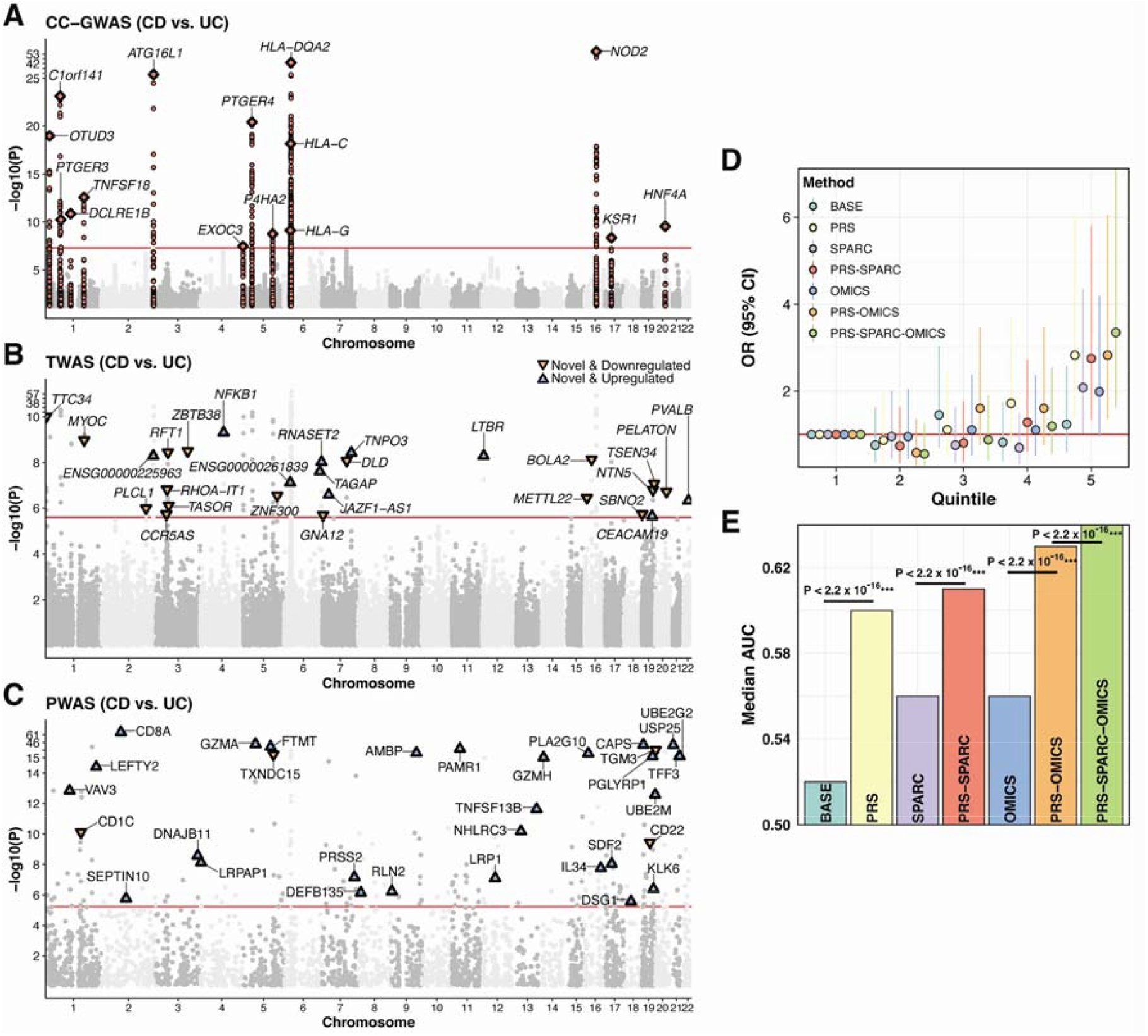
Integrative genetic, transcriptomic, and proteomic association analyses distinguishing Crohn’s disease (CD) from ulcerative colitis (UC). **(A)** Case-case genome-wide association study (GWAS) identifying loci associated with Crohn’s disease (CD) versus ulcerative colitis (UC). Diamonds indicate sentinel variants, and the red line marks the genome-wide significance threshold (P = 5 × 10^-8^). Two-sided P values were calculated using a chi-squared test with 1 degree of freedom. **(B)** Transcriptome-wide association study (TWAS) highlighting genes whose predicted expression distinguishes CD from UC. The red horizontal line represents the Bonferroni-corrected significance threshold (P = 2.5 × 10^-6^ for testing ∼20,000 genes). Upward-pointing triangles mark genes with increased expression in CD, while downward-pointing triangles indicate decreased expression. The most significant gene is labeled at each novel locus located outside a ±1 Mb window of GWAS sentinel variants. Two-sided P values were derived from TWAS Z scores using gene-based association tests. **(C)** Proteome-wide association study (PWAS) identifying proteins significantly associated with CD versus UC. The red horizontal line shows the Bonferroni-corrected significance threshold (P = 5.7 × 10^-6^ for 8,821 protein targets: 2,941 Olink targets and 5,880 SomaScan targets). Upward-pointing triangles denote proteins upregulated in CD, and downward-pointing triangles indicate proteins downregulated in CD. The most significant protein is labeled at each novel locus outside GWAS and TWAS signals, defined as ±1 Mb around sentinel variants or gene boundaries. Two-sided P values were derived from PWAS Z scores using protein-based association tests. **(D)** The odd ratios (OR) and 95% confidence intervals (CIs; bars) of each risk score stratus against individuals in the lowest quintile for multiple predictive models distinguishing CD from UC: baseline model (BASE), polygenic risk score (PRS), SPARC, PRS-SPARC, integrated omics (OMICS), PRS-OMICS, and combined PRS-SPARC-OMICS approaches. We stratified risk score into 5 groups with the bottom quintile used as the reference group. The red horizontal line indicates OR = 1. **(E)** Predictive performance of risk models distinguishing CD from UC, showing median area under the curve (AUC) for BASE, PRS, SPARC, PRS-SPARC, OMICS, PRS-OMICS, and PRS-SPARC-OMICS models. P values are derived from two-sided Mann-Whitney U test, comparing the accuracy of different prediction models.

Furthermore, these findings consistently highlight NF-*κ*B as one of the central hubs differentiating CD from UC. Specifically, *NOD2* mutations result in the inappropriate activation of NF-*κ*B signaling in monocytes^46^. Prostaglandin receptors *PTGER4* interacts directly with NF-*κ*B pathway and attenuates macrophage activation^47^. NF-*κ*B signaling is essential in generating and maintaining CD8^+^ T cell memory^48^. *IL34* is induced by NF-*κ*B signaling via TNF-*α* and able to induce human monocyte differentiation^49,50^. *PLA2G10* activity promotes arachidonic acid metabolism, linking phospholipase A2 signaling to NF-*κ*B–driven inflammatory cascades^51^. Finally, *TFF3* modulates NF-*κ*B in intestinal epithelial cells^52^. Together, these results converge to implicate NF-κB as one of the unifying regulators across multiple genetic layers (GWAS, TWAS, and PWAS), providing mechanistic insight into CD- vs. UC-specific pathogenesis.

### Integrating PRS with triangulated plasma protein levels led to an improvement in diagnostic accuracy

Next, we evaluated the utility of integrating PRS with proteomic data to improve the accuracy of distinguishing CD from UC. While previous studies have demonstrated the value of combining genetics, serum antibodies, and smoking history^53^, no study to date has assessed the utility of integrating genetic and proteomic data for this purpose. Using data from the UK Biobank, we identified 565 IBD patients (193 CD, 372 UC) and divided them into training and testing cohorts based on the timing of plasma protein measurements relative to diagnosis. Specifically, the training set included 87 CD and 171 UC cases, while the testing set included 106 CD and 201 UC cases. This cohort included 270 males (84 CD, 186 UC) and 295 females (109 CD, 186 UC), with year of birth (YOB) ranging from 1938 to 1969.

We depicted the performance of each model (**Figure 8D-E**) in stratifying individuals with CD vs. UC in the testing dataset. With the **BASE model** (sex, year of birth, and 20 principal components), there were 1.23 times as many CD cases in the top quintile as in the bottom quintile (OR 1.23, 95% CI [0.59–2.58]); the median area under the curve (AUC) was 0.52. Adding the polygenic risk score (**PRS model**) increased the AUC to 0.60 (P value < 2.2 × 10^-16^), with 2.82 times as many CD cases in the top quintile as in the bottom (OR 2.82, 95% CI [1.35–6.06]). Using 13 proteins from the SPARC IBD cohort^54^, the **SPARC model** achieved a median AUC of 0.56, with 2.08 times as many CD cases in the top quintile as in the bottom quintile (OR 2.08, 95% CI [1.02–4.34]); combining these proteins with PRS (**PRS–SPARC model**) raised the AUC to 0.61 (P value < 2.2 × 10^-16^), with 2.75 times as many CD cases in the top quintile as in the bottom quintile (OR 2.75, 95% CI [1.33–5.82]).

From our own analysis, leveraging TWAS and PWAS signals to triangulate for predictive differential regulators, we identified 8 proteins: AGER, APOM, ATP6V1G2, DDR1, HSPA1A, LTBR, RNASET2, and TNXB. Providing further validity to our approach, several of these proteins have been previously implicated in the core pathophysiology of IBD and its two subtypes. The AGER/RAGE signaling pathway has been implicated in driving intestinal inflammation in mouse models of IBD^55^. DDR1 has been shown to promote intestinal barrier disruption in UC^56^, and intestinal expression of HSPA1A has been noted to be elevated in patients with UC, but less so in CD or infectious colitis^57^. Polymorphisms in RNASET2 expression have been associated with CD severity and resistance to therapy^58^. Using these proteins (**OMICS model**) produced a median AUC of 0.56 and 1.99 times as many CD cases in the top quintile as in the bottom quintile (OR 1.99, 95% CI [0.96–4.20]); integrating these proteins with PRS (**PRS–OMICS model**) improved the AUC to 0.63 (P value < 2.2 × 10^-16^), with 2.82 times as many CD cases in the top quintile as in the bottom quintile (OR 2.82, 95% CI [1.36– 6.06]). Finally, the combined **PRS–SPARC–OMICS model** achieved the best performance, yielding a median AUC of 0.64 (P value < 2.2 × 10^-16^) and 3.35 times as many CD cases in the top quintile as in the bottom quintile (OR 3.35, 95% CI [1.62–7.14]). Importantly, germline genetic variants do not change over the lifetime and pQTL effects are derived to relatively healthy individuals. We hypothesize that the proteins identified through our multi-omics analyses will likely capture early molecular signals in otherwise healthy individuals. Together with the SPARC-derived proteins that likely capture transient-disease state activity, our approach offers complementary information that enhances prediction of new-onset CD and UC.

## DISCUSSION

In this study, we systematically characterized the genetic architecture of plasma proteins across two major proteomic platforms, including Olink and SomaScan, leveraging data from over 90,000 individuals. Our cross-platform comparison estimated a median genetic correlation of 0.49, signifying agreement between the two platforms but also highlighting substantial platform-specific signals. While both platforms captured well-known pleiotropic loci such as *HLA, ABO*, and *SH2B3*, we identified unique associations likely attributable to differences in target design, epitope recognition, and assay sensitivity.

Through meta-analysis and multi-trait approaches, we substantially increased the power of pQTL discovery and replication. MTAG improved power across both platforms, particularly for SomaScan, where the number of sentinel pQTLs was more than doubled. These gains directly translated to improved downstream analyses, including TWAS for protein levels, which identified >100K upstream regulators with strong cross-cohort replication. Tissue enrichment analyses further highlighted the liver as the principal contributor to the circulating proteome, consistent with prior studies noting its abundance of secreted proteins^59^. The small intestine, adipose tissue, spleen, and whole blood were also found to be major sources of the proteome, consistent with observations from the Human Protein Atlas^60^.

We generated a large GWAS dataset directly comparing CD and UC, representing the first effort of this scale^61^. We demonstrated the utility of integrating TWAS and PWAS to identify key protein regulators that distinguish the two subtypes, revealing multiple divergent loci enriched for NF-*κ*B signaling components. Importantly, incorporating proteomic features with PRS markedly enhanced CD vs. UC classification. Our findings suggest that proteins identified through our multi-omics approach (AGER, APOM, ATP6V1G2, DDR1, HSPA1A, LTBR, RNASET2, and TNXB) may represent more stable, genetically influenced features that better reflect healthy states, in contrast to the more transient disease-state markers likely captured in the SPARC cohort. We propose that combining these complementary marker sets could enable earlier and more reliable differentiation of CD from UC, thereby improving diagnostic precision and providing durable insights into IBD subtype pathogenesis if validated.

Our results add to a growing body of research leveraging omics-based approaches to improve disease discrimination and diagnosis. Recent studies have demonstrated the utility of protein-modification signatures^62^ and multi-omics-based machine learning models to distinguish UC from CD^63^. Building on these efforts, our integration of large-scale pQTL datasets, GWAS, and TWAS provides complementary mechanistic insights into how molecular regulation diverges between subtypes, with potential diagnostic relevance.

We acknowledge several limitations. First, the pQTL datasets analyzed were only derived from individuals of European ancestry, which limits the generalizability of our findings across diverse populations. Expanding analyses to include samples from underrepresented ancestries will be critical to ensure broader applicability and to capture ancestry-specific genetic effects. Second, while our study leveraged some of the largest available proteomic and genetic resources, the number of IBD cases with paired genomic and proteomic data is limited. Larger datasets with more diseased individuals will be needed to validate and refine the predictive models and to uncover additional subtype-specific regulators. Finally, our approach was cross-sectional in nature and did not allow for evaluation of longitudinal changes in the proteome across the lifespan.

Overall, our findings provide a comprehensive resource for plasma protein genetics and support the utility of integrative multi-omics approaches in disease biology and risk stratification. We can apply the framework to study other difficult-to-distinguish diseases, paving the way for more precise diagnostics and advancing the practice of precision medicine.

## METHODS

### Quality control of pQTL datasets in Olink and SomaScan platforms

We first compiled previously published pQTL datasets utilizing Olink and SomaScan platforms to quantify plasma protein levels. Specifically, for the Olink platform, we retrieved pQTL datasets from the UK Biobank^13^ and EPIC-Norfolk^3^ cohorts. For the SomaScan platform, we utilized pQTL datasets from deCODE^6^, AGES^8^, INTERVAL^5^, KORA^4^, and Fenland^7^ cohorts. We harmonized pQTL datasets to hg38 and calibrated the effect allele to the reference allele. We also ensured that participating studies have little or no sample overlaps and the effect sizes are homogeneous between studies of similar ancestry, by calculating the *λ*_*meta*_ statistic as previously described^64,65^. We limited our analysis to SNPs with MAF >1% in 503 European samples from the 1000 Genomes Project Phase 3^66^. For the discovery phase, we used the UK Biobank cohort for the Olink platform and used the deCODE, AGES, INTERVAL, and KORA cohorts for the SomaScan platform. For the replication phase, we utilized EPIC-Norfolk for the Olink platform and Fenland for the SomaScan platform.

### Meta-analysis of pQTLs across SomaScan studies and across platforms

Since there are multiple available SomaScan cohorts (deCODE, AGES, INTERVAL, and KORA), we first performed Z-score-based fixed-effect meta-analysis as implemented in METAL software^67^. Specifically, we matched each study from each SomaScan cohort via sequence ID (e.g., 10000-28). Next, we used LDSC software to calculate genetic correlations of the overlapped proteins via UniProt ID (e.g., P43320) between Olink and SomaScan platforms. For quality control, we compared the protein level correlation across platforms using individual-level data^13^, with our genetic correlation via LDSC. We considered traits to be genetically correlated if genetic correlation P values (two-sided) are significant after controlling false discovery rate (FDR) at 0.05 level. We then performed multi-trait analysis combining significantly positively correlated pQTLs using the MTAG software^23^. The HLA region, defined as chromosome 6 (25.5 Mb – 34 Mb), as well as *cis* region, defined as ±1 Mb window surrounding gene start or end, were excluded from the multi-trait meta-analysis due to its unusually large effect sizes that violate the model assumptions for MTAG.

### Definition of significant pQTL loci and replication analysis

Genome-wide significant threshold was defined as 5 × 10^-8^ / 8,821 (adjusted for 8,821 protein targets profiled across Olink and SomaScan platforms). Independent loci were defined as ±1 Mb window surrounding sentinel variants. Association was *cis* if the variant is within ±1 Mb of the transcription start and end sites. In contrast, *trans* association was defined as variants located outside a 1Mb window from transcription start and end sites and variants located on different chromosomes. We then annotated potential target genes for each sentinel variant according to the Open Target Genetics database, which used functional annotation data (e.g., eQTL, pQTL, and pc-HiC) to link regulatory variants to target genes^68^.

For pQTL replication analysis, we first identified variant-protein pairs in the discovery analysis with association P value < 5 × 10^-8^ / 8,821. To be replicated, the exact variant-protein pairs in the replication analysis must have an association P value < 0.05 and effect sizes in the same direction as the discovery sample.

### Tissue enrichment analysis

Using genome-wide pQTLs as input, we performed tissue enrichment analysis via LDSC-SEG^69^, following the same procedure as in GWAS enrichment analysis. Specifically, we utilized a precompiled set of tissue-specific genes from GTEx (https://github.com/bulik/ldsc/wiki/Cell-type-specific-analyses). Protein levels are considered enriched in a given tissue if the P values are significant after controlling FDR at the 0.05 level.

### TWAS analysis and replication analysis

We performed TWAS analysis using PUMICE software, which leverages epigenetic and 3D genomic information to more accurately create gene expression prediction models^27^. We created models based on genotype data and gene expression levels from GTEx v8 to impute models in 49 different human tissues^70^. Only *cis*-eQTL with MAF >0.01 were included to build these models. For each tissue, we conducted TWAS of each protein target and considered transcript-protein pair with P value < 0.05 / (20,000 x 8,821) to be statistically significant (adjusted for 20,000 transcripts and 8,821 protein targets). We defined *cis*-same, *cis*-different, and *trans*-acting gene regulators of protein level as previously described^29^. *Cis*-same associations were defined as transcripts linked to the protein encoded by the same gene. *Cis*-different associations were defined as transcripts linked to the protein encoded by a different gene within ±1 Mb of the transcript boundaries. *Trans* associations were defined as transcripts linked to proteins encoded by genes located more than 1 Mb away or on different chromosomes.

For TWAS replication analysis, we first identified transcript-protein pairs in the discovery analysis with association P value < 5 × 10^-8^ / (20,000 x 8,821). Given each significant transcript-protein pair may exist in multiple tissues, we only retained the most significant association. To be considered a successful replication, the exact transcript-protein pairs in the replication analysis need to have association P value < 0.05 and the same direction of effect size.

### Protein level prediction models

We created protein level prediction models using PRSCS^42^ with default settings. Specifically, we used genome-wide summary-statistic pQTLs of each protein along with 1000 Genomes linkage disequilibrium reference panel as inputs to PRSCS software. The model was considered significant if correlation coefficient was greater than 0.1 and estimated P value was less than 0.05 when evaluated in the replication cohorts.

We do not have individual level protein abundance and genotype data to validate prediction accuracy. Instead, we rely on pQTL summary statistics from an independent study to assess the accuracy approximately. We use the following formula to calculate the correlation between measured and predicted protein abundance:

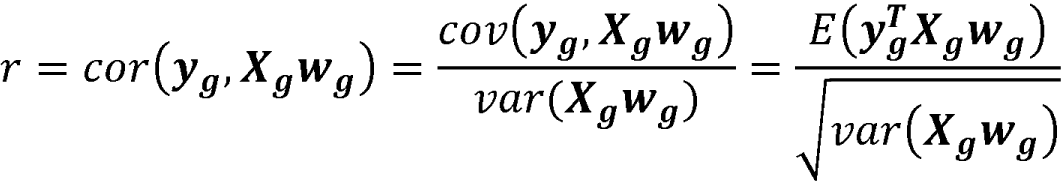

Where ***y***_***g***_ is the measured protein level and ***X***_***g***_ is the genotype from the prediction cohort. Notably, 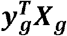 can be approximated by the external pQTL effect sizes and ***X***_***g***_***w***_***g***_ could also be derived from the reference LD panel. So *r* can still be calculated if only pQTL summary statistics are available from the test data.

To calculate the p-value of the derived correlation, we use Fisher’s transformation to obtain the z-score from correlation coefficients:

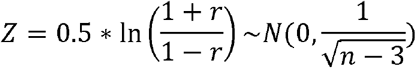

The converted Z-score follows a normal distribution. The prediction model is deemed significant if *r* > 0.1 and p-value < 0.05, following the standard procedure in PWAS^27^.

### GWAS of CD vs. UC

We downloaded two distinct publicly available GWAS datasets from GWAS catalog: 1) CD vs. Healthy controls GWAS (12,194 cases vs. 28,072 controls) and 2) UC vs. Healthy controls GWAS (12,366 cases vs. 33,609 controls)^71,72^. These two GWAS datasets were used to create CD vs. UC GWAS dataset using CC-GWAS^43^, which tests for differences in allele frequency between cases of two disorders using summary statistics from the respective case-control GWAS. The prevalence of CD and UC were estimated as 0.3% (95% CI 0.1%, 0.4%) and 1.0% (95% CI 0.5%, 1.4%), respectively^73^. Using stratified LD Score regression, the estimated heritability on a liability scale for CD and UC were 0.27 and 0.22, respectively. Using cross-trait LD Score regression, the estimated genetic correlation between CD and UC was 0.63 and intercept was 0.29. We specified approximation of number of independent effective loci (m) as 1,000. Genome-wide significant threshold was defined as 5× 10^-8^.

### Genetic imputation of transcriptome and proteome in CD vs. UC

We performed TWAS using gene expression prediction models across 49 GTEx tissues constructed via PUMICE and considered transcripts to be significantly associated with CD vs. UC trait if P values were less than 0.05/20,000. We then performed PWAS using protein level prediction models created via PRS-CS and considered proteins to be significantly associated with CD vs. UC trait if P values were less than 0.05/8,821, which is the Bonferroni threshold correcting for the number of tested proteins. Independent loci for both TWAS and PWAS were defined as ±1 Mb window surrounding transcript start and end sites.

### UK Biobank dataset

We obtained individual-level clinical, genetic, and proteomic data under application number 21237. For genetics data, we subsetted for 361,549 European individuals and 13,791,467 variants previously used in UKBB GWAS imputed V3 (http://www.nealelab.is/uk-biobank/). For proteomic data, we retrieved the levels of protein measured in 2,923 proteins across 53,014 individuals. In summary, there were 38,334 European individuals who have both genetic and proteomic data available for downstream analysis. To identify patients with CD and UC, we searched for ICD-9 and ICD-10 codes [CD: ICD-9 555, ICD-10 K50; UC: ICD-9 556, ICD-10 K51] in fields 41270-41271 and associated first diagnosis dates in fields 41280-41281. In order to determine date of blood collection, the information in field 53 was used. We excluded patient who had both CD and UC diagnoses. We subsequently defined patients who had their blood collected after disease diagnosis as prevalent cases, while patients who had their blood collected prior the diagnosis as incident cases. We used prevalent cases as our training dataset, while using incident cases as our testing dataset as this reflects the most realistic and practical application of biomarkers.

### Evaluation of prediction performance

Specifically, we performed five-fold cross-validation using elastic net regression (*α* = 0.5) in the training set and generated predictions in the testing set. To account for the stochastic nature of the algorithm, cross-validation was repeated 50 times with different random seeds (1–50). Odds ratios for CD versus UC were calculated by comparing each risk-score quintile with the bottom 20th percentile (seed = 1). For area under the receiver operating characteristic curve (AUC), we reported the median AUC across runs (seed 1-50). Pairwise comparisons of AUCs between models were conducted using the two-sided Mann–Whitney U test.

### Software URLs

CC-GWAS software can be found at https://github.com/wouterpeyrot/CCGWAS/.

LDSC software can be found at https://github.com/bulik/ldsc.

MTAG software can be found at https://github.com/JonJala/mtag.

Open Target Genetics software can be found at https://genetics.opentargets.org.

PLINK software can be found at https://www.cog-genomics.org/plink.

PRS-CS-auto software can be found at https://github.com/getian107/PRScs.

PUMICE software can be found at https://github.com/ckhunsr1/PUMICE.

### Data availability

The pQTL summary statistics of the SomaScan meta-analysis and cross-platform meta-analysis, along with TWAS results and PWAS models, will be deposited in a Shiny App, where users can download the data and interactively explore the findings upon publication. We obtained UKB-PPP pQTL summary statistics from https://www.decode.com/summarydata/. We obtained EPIC-Norfolk pQTL summary statistics from https://omicscience.org/. We obtained deCODE pQTL summary statistics from https://www.decode.com/summarydata/. We obtained AGES pQTL summary statistics from https://www.ebi.ac.uk/gwas/home. We obtained INTERVAL pQTL summary statistics from http://www.phpc.cam.ac.uk/ceu/proteins/. We obtained KORA pQTL summary statistics from https://www.ebi.ac.uk/gwas/home. We obtained FENLAND pQTL summary statistics from https://omicscience.org/. We obtained CD and UC GWAS summary statistics from https://www.ebi.ac.uk/gwas/home.

## ACKNOWLEDGMENT

This work was supported by the National Institutes of Health grants R01HG011035, R01AI174108, R01ES036042, R01HL173869, and U01AI185638. This research has been conducted using the UK Biobank Resource under application number 21237. We thank M. Koprulu for providing us with EPIC-Norfolk data. We thank W. Peyrot for help with CC-GWAS method and implementation.

## AUTHOR CONTRIBUTIONS

C.K. and D.L. conceived the study. C.K. led the data analysis. C.K., F.Z., L.W., S.C., H.M., D.C. conducted analyses. G.S., C.L.S., F.J., A.K., X.Z., J.M., K.H., and B.J. helped with data interpretation. C.K. and D.L. prepared the manuscript. All authors contributed to manuscript editing and approved the manuscript.

